# IgG *N-*glycans are associated with prevalent and incident complications of type 2 diabetes

**DOI:** 10.1101/2022.03.15.22272417

**Authors:** Elham Memarian, Ralph Heijmans, Roderick C. Slieker, Adriana Sierra, Olga Gornik, Joline WJ Beulens, Maja Hanic, Petra Elders, Julio Pascual, Eric Sijbrands, Gordan Lauc, Viktoria Dotz, Clara Barrios, Leen M’t Hart, Manfred Wuhrer, Mandy van Hoek

## Abstract

**Aims/hypothesis:** Inflammation is important in development of type 2 diabetes complications. The *N*-glycosylation of IgG influences its role in inflammation. Until now, the association of IgG *N*-glycosylation with type 2 diabetes complications has not been extensively investigated. We hypothesized that *N*-glycosylation of IgG may be related to development of complications of type 2 diabetes.

**Methods:** In three independent type 2 diabetes cohorts, IgG *N*-glycosylation was measured by UPLC (DiaGene n=1815, GenodiabMar n=640) and mass spectrometry (DCS n=1266). We investigated the associations of IgG *N*-glycosylation (fucosylation, galactosylation, sialylation and bisection) with incident and prevalent nephropathy, retinopathy and macrovascular disease using Cox- and logistic regression, followed by meta-analyses. The models were adjusted for age, sex and additionally for clinical risk factors.

**Results:** IgG galactosylation was negatively associated with prevalent and incident nephropathy after adjustment for clinical risk factors. Sialylation was negatively associated with incident diabetic nephropathy. For retinopathy, similar associations were found for galactosylation in the basic model. For macrovascular complications, negative associations with galactosylation and sialylation were confined to the cross-sectional analyses.

**Conclusions:** We showed that IgG *N*-glycosylation traits are associated with higher prevalence and future development of nephropathy, after correction for clinical risk factors. For other complications, IgG *N*-glycosylation was associated with their prevalence only, possibly reflecting ongoing vascular inflammation. These findings indicate the predictive potential of IgG *N*-glycosylation in nephropathy.

## Introduction

Type 2 diabetes is one of the most common chronic diseases worldwide. The long term microvascular and macrovascular complications cause an enormous burden of the disease ^1^. It has been demonstrated that inflammation plays a key role in the pathophysiology of type 2 diabetes and its complications ^2^. Consequently, *N*-glycosylation of IgG might contribute to pathophysiological processes of type 2 diabetes complications as glycosylation is known to influence IgG effector functions ^3^. We therefore hypothesized that *N*-glycosylation of IgG may be involved in the development of complications of type 2 diabetes.

Posttranscriptional modifications of proteins are thought to have a role in almost every major disease. One of the most frequently-occurring posttranscriptional protein modifications is glycosylation ^4^. IgG *N*-glycosylation patterns vary between individuals, and contain a genetic, heritable component ^5^. However, IgG glycans show excellent temporal stability in a single individual and therefore have diagnostic potential ^6^. Changes in IgG *N*-glycome composition are associated with a variety of diseases such as rheumatoid arthritis, inflammatory bowel disease, cancer, kidney disease and type 2 diabetes ^7–9^. Recent retrospective studies have also shown associations between IgG glycosylation and diabetic retinopathy ^10^, ischemic stroke ^11^ and CVD risk score ^12^. In addition, our team has recently found relationships between *N*-glycosylation of total plasma proteins and type 2 diabetes complications ^13^. However, to date, the effects of IgG glycosylation on clinical outcome in type 2 diabetes patients have not been fully investigated. In the present study, we determined the relationships between IgG *N*-glycans and diabetic nephropathy, retinopathy and macrovascular complications in three large European prospective cohort studies. The outcomes can provide more insights into diabetes complication pathophysiology, could contribute to biomarker development and identify patients with a higher risk profile for developing complications associated with type 2 diabetes.

## Materials and methods

### Patient cohorts

Three cohorts were included in this study. The DiaGene study has been described in more detail elsewhere ^14^. In short, the DiaGene study is a multi-center all-lines of care prospective cohort study. Between 2006 and 2011, individuals with and without type 2 diabetes were approached for participation in the region of Eindhoven, the Netherlands. Only cases with type 2 diabetes (n=1886) were included in the current study.

The Hoorn Diabetes Care System (Hoorn DCS) is a cohort that is comprised of currently over 14,000 individuals with type 2 diabetes from the region of West Friesland in the Netherlands ^15^. Annual examinations data and biobanking materials for around 5500 persons, with the agreement to participate in DCS biobanks, have been collected. For this study, we randomly selected a subset of plasma samples from 1600 subjects who donated a sample in 2008/2009.

The GenoDiabMar study is a type 2 diabetes adult registry, recruited between 2012 and 2015 from the healthcare area Litoral-Mar of Barcelona, Spain ^16,17^. Patients older than 45 years with a medical history of type 2 diabetes for more than 10 years and treated with anti-diabetic drugs were included (n = 640 for the current study).

The medical ethics committees of the Erasmus Medical Center, VU University Medical Center Amsterdam, and Institut Mar d’Investigacions Mediques have approved DiaGene (DiaGene MEC 2004-230), Hoorn DCS (Hoorn DCS MEC 2007-57), and GenoDiabMar (MEC 2014/4323) studies, respectively. All individuals gave their written informed consent.

### Measurements and definitions of macro-and microvascular complications

Definitions for macro- and microvascular complications have been described in detail for DiaGene, GenoDiabMar and DCS cohorts ^14,15,17^. In short, retinopathy for all studies was scored by an ophthalmologist based on fundus photography, and for the current study defined as present or absent, where present includes all gradations and stages of diabetic retinopathy. Nephropathy was defined as microalbuminuria [albumin/creatinine-ratio (ACR) ≥2.5 mg/mmol for men or ≥3.5 mg/mmol for women] present at two of three consecutive measurements, or when high micro-albuminuria or macro-albuminuria was present at one measurement (ACR ≥12.5 mg/mmol for men or ≥17.5 mg/mmol for women) ^14,15,17^. Cardiovascular disease derived from medical records and questionnaires was defined as myocardial infarction, percutaneous coronary intervention/coronary artery bypass graft (PCI/CABG), cerebrovascular accident, transient ischemic attack, and peripheral arterial disease (PAD). Ischemic brain disease (IBD) comprised both cerebrovascular incidents and transient ischemic attacks combined.

### IgG N-glycosylation analysis and data quality control

IgG *N*-glycan isolation, release and labeling in the DiaGene and GenoDiabMar studies followed the same procedure and were described elsewhere ^9^. Briefly, IgG was isolated from 100 μL of plasma per sample on protein G monolithic plate (BIA Separations, Ajdovščina, Slovenia). Consequently, IgG *N*-glycans were released by PNGase F, fluorescently labeled with 2-aminobenzamide and cleaned up by hydrophilic interaction liquid chromatography solid phase extraction (HILIC-SPE) from the excess of reagents. IgG *N*-glycan profiles were obtained on Acquity UPLC instrument (Waters, Milford, MA, USA) and separated into 24 glycan peaks. The relative amount of glycans in each peak was expressed as a percentage (%) of the total integrated area. In Supplementary Table 1, a detailed description of the 24 glycan peaks is given. Out of 1886 participants with type 2 diabetes in the DiaGene study, plasma of 1837 participants was available. As 29 samples failed data quality control, a total of 1815 participants were included. In GenoDiabMar, data from 640 participants were used for statistical analysis after quality control.

Samples from the Hoorn DCS cohort were analysed using an ultrahigh resolution mass spectometry method which is described in detail in the Supplementary Method 1. In short, after capturing IgG from plasma samples, following enzymatic *N*-glycan release, sialic acids were stabilized by derivatization. Subsequently, hydrophilic interaction liquid chromatography (HILIC) solid phase extraction and sample spotting were performed, and IgG *N*-glycans were measured by matrix-assisted laser desorption/ionization Fourier-transform ion cyclotron resonance (MALDI-FTICR) MS ^18^. After performing quality control on mass spectra in the Hoorn DCS cohort, low intensity spectra were excluded, and 35 IgG glycan peaks were quantified. 19 glycan compositions were matched with the 24 UPLC glycan peaks and only the matching 19 peaks were used for further statistical analysis (Supplementary Table 1).

In all three cohorts, four IgG glycosylation derived traits were calculated for glycans that share structural similarities: fucosylation, galactosylation, sialylation, and bisecting GlcNAc as indicated in Supplementary Table 2. These features were used in the main analyses, with subsequent further detailed analyses in the 19 direct glycan peaks which were matched between the three cohorts.

### Statistical analyses

In baseline characteristics of the cohorts, continuous variables are expressed as mean and their standard deviations. Categorical variables are expressed as percentages.

Batch correction for glycan relative abundance was performed in all three cohorts. Initially, normalization of IgG glycans was performed using the total area under the expression peaks. After log-transformation of the relative abundance values, batch effects were corrected using the ComBat function in the R package *sva*, modified to correct for outliers.

Both baseline and prospective analyses of the IgG glycome were adjusted for covariates known to influence vascular complications and/or IgG glycosylation. We considered two models: model 1 adjusted for age, sex and their interaction. Age and sex are known to influence glycans, especially glycosylation profiles of females, with the biggest effect around menopausal age ^19^. Model 2 was additionally adjusted for BMI, HDL-cholesterol, non-HDL-cholesterol, hypertension, HbA1c, duration of diabetes and smoking. Missing data of clinical variables in model 2 were replaced with substituted values using imputation in all three cohorts. Associations between prevalence and incidence of macro- and microvascular complications at baseline and follow-up and the 4 main IgG glycosylation features (fucosylation, galactosylation, sialylation and bisection) were evaluated using logistic and Cox regression models, respectively. Similarly, as a secondary analysis, associations between the 19 IgG glycan peaks and vascular complications were assessed. The presence of a vascular complication was entered as dependent variable, glycosylation feature and adjusting covariates were entered as independent variables. For the prospective analyses individuals with prevalent events at baseline were not included. Time-to-event was defined as the time between date of inclusion and date of the complication during follow-up, date of death or censoring at the end of follow-up. Across cohorts, results were random-effects meta-analysed using the metagen function from the R package meta. The Benjamini-Hochberg procedure was used to correct for multiple testing and a FDR adjusted P-value < 0.05 was considered significant. For statistical analyses, we used IBM SPSS Statistics version 25 in the DiaGene study, STATA version 15.1 in the GenoDiabMar study, and R (version R-3.6.3) and RStudio (version 1.4.1106) in the Hoorn DCS cohort.

## Results

### Baseline characteristics

The clinical characteristics of the three study cohorts at baseline are shown in Table 1. All cohorts had a higher percentage of male participants, and the average age of patients with type 2 diabetes was approximately 66 years. The mean diabetes duration in the DiaGene, Hoorn DCS, and GenoDiabMar cohorts was 10.0 (SD=8.4), 7.1 (±5.8), and 16.8 (±8.8) years, respectively. The mean duration of follow-up in these cohorts was 7.0 (SD= ±2.1), 7.5 (SD= ±2.3), and 4.7 (SD= ±1.3) years. In all cohorts, the most prevalent microvascular and macrovascular complications were nephropathy and ischemic heart disease (IHD), respectively.

**Table 1.** DiaGene, Hoorn DCS, and GenoDiabMar cohort characteristics. Details on the cohorts including definitions and biochemical measurements have been described elsewhere (12–14). For complications, numbers of participants are shown, and percentages are presented as n-case divided by n-total. (SD is defined as Standard deviation).

|  | **DiaGene** | **Hoorn DCS** | **GenoDiabMar** |
| --- | --- | --- | --- |
| Number of participants, n | 1815 | 1266 | 640 |
| Sex, n (% male) | 975 (53.7) | 706 (55.8) | 389 (60.8) |
| Age in years, mean (±SD) | 65.2 (±10.5) | 64.6 (±10.6) | 69.65 (±9.3) |
| Duration of diabetes (years), mean (±SD) | 10.0 (±8.4) | 7.1 (±5.8) | 16.82 (±8.8) |
| Duration of follow-up (years), mean (±SD) | 7.0 (±2.1) | 7.5 (±2.3) | 4.7 (±1.3) |
| BMI<sup>†</sup>, kg/m<sup>2</sup>, mean (±SD) | 30.5 (±5.5) | 30.4 (±5.4) | 30.31 (±5.1) |
| HbA1c<sup>‡</sup>, %, mean (±SD) | 7.03 (±1.06) | 6.78 (±1.0) | 7.72 (±1.3) |
| HbA1c, mmol/mol, mean | 53 | 51 | 61 |
| Mean Arterial Pressure (mmHg), (±SD) | 98.9 (±10.8) | 99.0 (±11.3) | 95.10 (±11.4) |
| HDL-c<sup>□</sup>, mmol/L, mean (±SD) | 1.17 (±0.3) | 1.18 (±0.4) | 1.22 (±0.3) |
| Non-HDL-c<sup>£</sup>, mmol/L, mean (±SD) | 3.12 (±0.9) | 3.46 (±1.5) | 3.27 (±0.9) |
| **Smoking** |  |  |  |
| Never, n (%) | 602 (33.2) | 783 (61.9) | 259 (43.6) |
| Former, n (%) | 921 (50.7) | 269 (21.2) | 222 (37.4) |
| Current, n (%) | 292 (16.1) | 214 (16.9) | 113 (17.8) |
| **Complications** |  |  |  |
| Prevalent IHD<sup>¶</sup>, n (%) | 467 (25.7) | 199 (15.7) | 134 (20.9) |
| Incident IHD, n (%) (n-case/n-total) | 45 (3.6) | 71 (5.6) | 74 (11.6) |
| Prevalent PAD<sup>§</sup>, n (%) (n-case/n-total) | 179 (9.9) | 35 (2.8) | 125 (19.5) |
| Incident PAD, n (%) (n-case/n-total) | 93 (6.1) | 17 (1.3) | 54 (8.4) |
| Prevalent IBD<sup>¥</sup>, n (%) (n-case/n-total) | 200 (11.0) | 29 (2.3) | 67 (10.5) |
| Incident IBD, n (%) (n-case/n-total) | 63 (4.2) | 14 (1.1) | 28 (4.4) |
| Prevalent Nephropathy, n (%) (n-case/n-total) | 352 (19.4) | 199 (15.7) | 347 (54.2) |
| Incident Nephropathy, n (%) (n-case/n-total) | 243 (20.0) | 131 (10.3) | 129 (20.2) |
| Prevalent Retinopathy, n (%) (n-case/n-total) | 288 (15.9) | 172 (13.7) | 162 (25.3) |
| Incident Retinopathy, n (%) (n-case/n-total) | 220 (16.2) | 104 (8.2) | 134 (20.9) |

<sup>†</sup> BMI: Body mass index
<sup>‡</sup> HbA1c: Hemoglobin A1C
<sup>□</sup> HDL-c: High-density lipoprotein (HDL) cholesterol
<sup>£</sup> Non-HDL-c, Cholesterol value minus HDL cholesterol
<sup>¶</sup> IHD: Ischemic Heart Disease
<sup>§</sup> PAD: Peripheral Artery Disease
<sup>¥</sup> IBD: Ischemic Brain Disease

IgG main glycan features that were significantly associated with microvascular and macrovascular complications after FDR correction at baseline and during follow-up in the meta-analysis are shown highlighted in Tables 2 and 3. Several nominally significant results were found that did not remain significant after FDR correction, they are shown in table 2 and 3 in bold without highlight.

**Table 2.** Meta-analyzed associations of main IgG glycosylation features with type 2 diabetes complications. Nominal-significant values are marked bold (*p* < 0.05), and values highlighted in yellow are FDR-significant (Basic model: Age, sex, age×sex interaction). (OR and HR are defined as odds ratio, and hazard ratio, respectively).

|  | Nephropathy |  |  |  | Retinopathy |  |  |  | IHD<sup>†</sup> |  |  |  | PAD<sup>‡</sup> |  |  |  | IBD<sup>□</sup> |  |  |  |
| --- | --- | --- | --- | --- | --- | --- | --- | --- | --- | --- | --- | --- | --- | --- | --- | --- | --- | --- | --- | --- |
|  | Prevalent (Logistic) |  | Incident (Cox) |  | Prevalent (Logistic) |  | Incident (Cox) |  | Prevalent (Logistic) |  | Incident (Cox) |  | Prevalent (Logistic) |  | Incident (Cox) |  | Prevalent (Logistic) |  | Incident (Cox) |  |
| Derived traits | OR | *p*-value | HR | *p*-value | OR | *p*-value | HR | *p*-value | OR | *p*-value | HR | *p*-value | OR | *p*-value | HR | *p*-value | OR | *p*-value | HR | *p*-value |
| Galactosylation | **0.74** | **8.08x10<sup>-6</sup>** | **0.81** | **3.68x10<sup>-5</sup>** | **0.86** | **1.99x10<sup>-3</sup>** | **0.86** | **1.05x10<sup>-2</sup>** | **0.88** | **6.23x10<sup>-3</sup>** | 0.72 | 0.06 | **0.84** | **3.67x10<sup>-2</sup>** | 0.83 | 0.06 | 0.89 | 0.09 | 0.91 | 0.41 |
| Bisection | 0.89 | 0.18 | 0.94 | 0.17 | **0.86** | **5.98x10<sup>-4</sup>** | 0.97 | 0.55 | 1.1 | 0.11 | **0.84** | **3.38x10<sup>-2</sup>** | 1.09 | 0.51 | 1.09 | 0.37 | 1 | 0.99 | 1.03 | 0.76 |
| Sialylation | **0.86** | **3.57x10<sup>-2</sup>** | **0.83** | **5.89x10<sup>-3</sup>** | 0.96 | 0.41 | 0.97 | 0.56 | **0.88** | **5.58x10<sup>-3</sup>** | 0.85 | 0.40 | 0.89 | 0.26 | 0.87 | 0.15 | 0.89 | 0.08 | 0.94 | 0.58 |
| Fucosylation | 0.98 | 0.67 | 1.01 | 0.93 | 1.02 | 0.59 | 0.97 | 0.51 | 0.96 | 0.29 | 0.92 | 0.44 | 0.92 | 0.12 | 0.99 | 0.96 | 1 | 0.95 | 1.06 | 0.67 |

<sup>†</sup> IHD: Ischemic Heart Disease
<sup>‡</sup> PAD: Peripheral Artery Disease
<sup>□</sup> IBD: Ischemic Brain Disease

**Table 3.** Meta-analyzed associations of main IgG glycosylation features with type 2 diabetes complications. Nominal-significant values are marked bold (*p* < 0.05), and values highlighted in yellow are FDR-significant (Full model: Age, sex, age×sex interaction, BMI, HDL-cholesterol, non-HDL-cholesterol, hypertension, HbA1c, duration of diabetes and smoking) (OR and HR are defined as odds ratio, and hazard ratio, respectively).

| Derived traits | Nephropathy |  |  |  | Retinopathy |  |  |  | IHD<sup>†</sup> |  |  |  | PAD<sup>‡</sup> |  |  |  | IBD<sup>□</sup> |  |  |  |
| --- | --- | --- | --- | --- | --- | --- | --- | --- | --- | --- | --- | --- | --- | --- | --- | --- | --- | --- | --- | --- |
|  | Prevalent (Logistic) |  | Incident (Cox) |  | Prevalent (Logistic) |  | Incident (Cox) |  | Prevalent (Logistic) |  | Incident (Cox) |  | Prevalent (Logistic) |  | Incident (Cox) |  | Prevalent (Logistic) |  | Incident (Cox) |  |
|  | OR | *p*-value | HR | *p*-value | OR | *p*-value | HR | *p*-value | OR | *p*-value | HR | *p*-value | OR | *p*-value | HR | *p*-value | OR | *p*-value | HR | *p*-value |
| Galactosylation | **0.79** | **3.03x10<sup>-3</sup>** | **0.86** | **3.67x10<sup>-3</sup>** | **0.88** | **1.79x10<sup>-2</sup>** | 0.9 | 0.09 | 0.93 | 0.15 | 0.73 | 0.10 | **0.83** | **1.09x10<sup>-2</sup>** | 0.86 | 0.13 | 0.93 | 0.295 | 0.9 | 0.41 |
| Bisection | 0.92 | 0.31 | 0.95 | 0.25 | **0.89** | **1.67x10<sup>-2</sup>** | 1.01 | 0.88 | **1.13** | **1.80x10<sup>-2</sup>** | 0.85 | 0.06 | 1.08 | 0.61 | 1.03 | 0.74 | 0.99 | 0.964 | 1.04 | 0.73 |
| Sialylation | 0.91 | 0.14 | **0.86** | **4.88x10<sup>-3</sup>** | 0.96 | 0.48 | 0.98 | 0.69 | 0.93 | 0.12 | 0.85 | 0.42 | 0.89 | 0.13 | 0.9 | 0.28 | 0.94 | 0.361 | 0.95 | 0.63 |
| Fucosylation | 0.98 | 0.65 | 1.02 | 0.77 | 1.01 | 0.87 | 0.96 | 0.39 | 0.93 | 0.08 | 0.91 | 0.32 | 0.95 | 0.35 | 1 | 0.10 | 1.03 | 0.661 | 1.11 | 0.61 |

<sup>†</sup> IHD: Ischemic Heart Disease
<sup>‡</sup> PAD: Peripheral Artery Disease
<sup>□</sup> IBD: Ischemic Brain Disease

A graphical representation of all significant results and total results can be found in Figure 1 and Supplementary Figure 1, respectively. Associations between the individual 19 IgG *N*-glycan peaks and microvascular and macrovascular complications can be found in Supplementary Table 3. Heterogeneity between the three cohorts was observed (Supplementary Table 3). The level of heterogeneity can be explained by differences in cohort backgrounds and clinical data collection.

**Figure 1:**
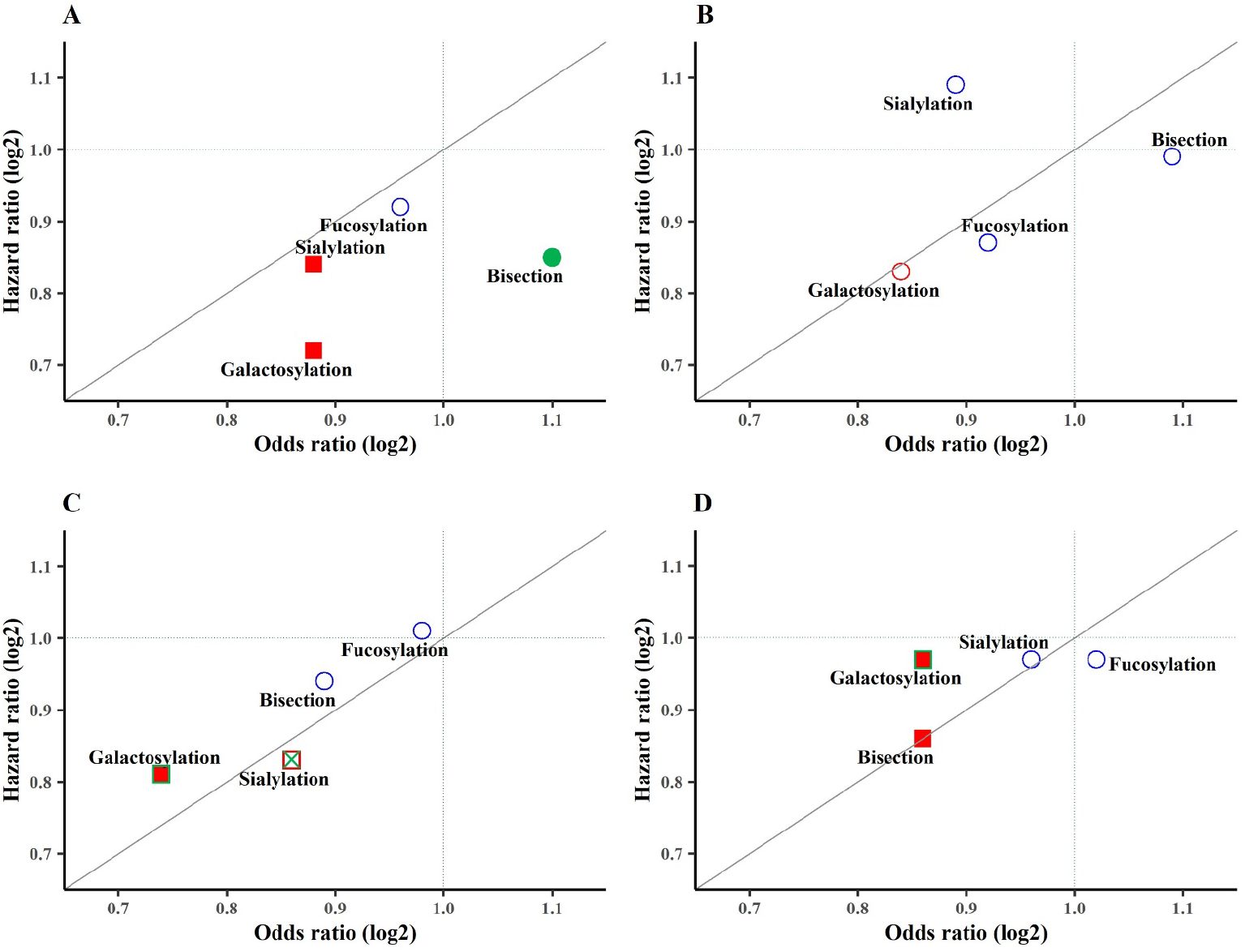
*N*-glycan derived traits odds ratios (OR) and hazard ratios (HR) for meta-analyzed data from DiaGene, Hoorn DCS, and GenoDiabMar studies in the basic model (adjusted for age, sex, and age×sex interaction). (A) Ischemic Heart Disease, (B) Peripheral Artery Disease, (C) Nephropathy, (D) Retinopathy. Red-filled green square: Significant in prevalent and incident complications after FDR correction. Red unfilled square with green cross: Significant in prevalent complications before FDR correction and in incident complications after FDR correction. Red-filled square: Significant in prevalent complications after FDR correction. Green-filled circle: Significant in incident complications before FDR correction. Red-filled circle with green cross: Significant in incident complications after FDR correction. Red unfilled circle: Significant in prevalent complications before FDR correction. Blue unfilled circle: Non-significant.

### Microvascular complications

In the meta-analysis, galactosylation was negatively associated with prevalent (OR = 0.74, 95% CI = 0.65-0.84, *p*_FDR_ = 3.23×10^−5^) as well as incident diabetic nephropathy (HR = 0.81, 95% CI = 0.73-0.89, *p*_FDR_ = 1.47×10^−4^), in the basic model and this remained significant after adjustment for clinical variables in model 2 (OR = 0.79, 95% CI = 0.68-0.93, *p*_FDR_ = 1.21×10^−2^ and HR = 0.86, 95% CI = 0.77-0.95, *p*_FDR_ = 1.46×10^−2^). In addition, sialylation was negatively associated with incident diabetic nephropathy in both models (model 1; HR = 0.83, 95% CI = 0.73-0.95, *p*_FDR_ = 1.77×10^−2^ and model 2; HR = 0.86, 95% CI = 0.78-0.96, *p*_FDR_ = 1.46×10^−2^) (Supplementary Table 4). Figure 2 shows the effect sizes across all cohorts for galactosylation and sialylation regarding diabetic nephropathy. In Supplementary Figures 2-4 effect sizes across all cohorts can be found for all vascular complications. Considering the 19 IgG *N*-glycan peaks, the association with nephropathy appeared to be mainly driven by 2 glycan peaks. The strongest, statistically significant, negative associations in the basic model were found for digalactosylated (GP14) and monosialylated digalactosylated fucosylated biantennary glycans (GP18) in the prospective analyses (HR = 0.81, 95% CI = 0.72-0.91, *p*_FDR_ = 2.57×10^−3^ and HR = 0.81, 95% CI = 0.71-0.93, *p*_FDR_ = 1.41×10^−2^, respectively) (Supplementary Table 3).

**Figure 2.**
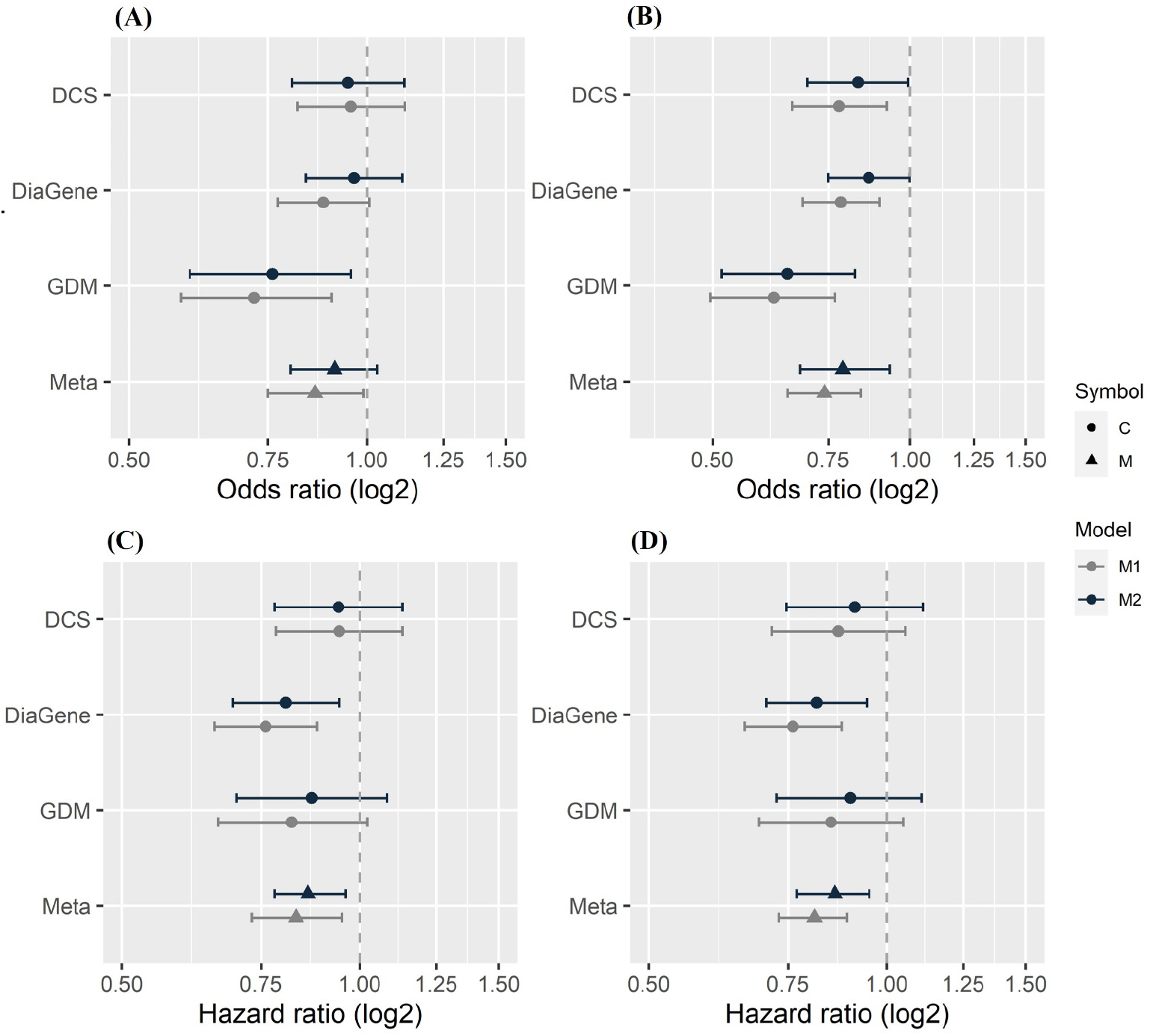
Forest plots of associations of *N*-glycan sialylation and galactosylation derived traits with nephropathy. Data from DiaGene, Hoorn DCS, and GenoDiabMar (GDM) studies and meta-analyzed data are from the basic and full model analysis. (A) IgG Sialylation in Nephropathy (prevalent), (B) IgG Galactosylation in Nephropathy (prevalent), (C) IgG Sialylation in Nephropathy (incident), (D) IgG Galactosylation in Nephropathy (incident).

Galactosylation was negatively associated with diabetic retinopathy, in both cross-sectional (OR = 0.86, 95% CI = 0.78-0.95, *p*_FDR_ = 5.96×10^−3^) and prospective analyses (HR = 0.86, 95% CI = 0.77-0.97, *p*_FDR_ = 4.18×10^−2^). However, after adjustment for clinical confounders, these associations did not remain statistically significant. A negative association between bisection and diabetic retinopathy prevalence was found in the basic model (OR = 0.86, 95% CI = 0.79-0.94, *p*_FDR_ = 2.39×10^−3^).

### Macrovascular complications

Regarding ischemic heart disease (IHD), a negative association with galactosylation (OR = 0.88, 95% CI = 0.80-0.96, *p*_FDR_ = 1.87×10^−2^) and sialylation (OR = 0.88, 95% CI = 0.81-0.96, *p*_FDR_ = 1.87×10^−2^) was found in the basic model at baseline. No significant prospective associations were found.

Concerning peripheral artery disease (PAD), a negative association between galactosylation and prevalent PAD was found, after adjustment for clinical confounders (OR = 0.83, 95% CI = 0.73-0.96, *p*_FDR_ = 4.37×10^−2^). These results appear to be mainly driven by digalactosylated (GP14) and disialylated digalactosylated fucosylated biantennary glycans (GP23), which were significantly associated with lower PAD and IHD prevalence in the full model, respectively (OR = 0.77, 95% CI = 0.66-0.89, *p*_FDR_ = 8.84×10^−3^ and OR = 0.85, 95% CI = 0.75-0.95, *p*_FDR_ = 3.58×10^−2^, respectively) (Supplementary Table 3). No prospective associations were observed. Ischemic brain disease (IBD) was not significantly associated with the four main glycan features nor with any of the 19 direct glycan peaks (Supplementary Table 3,4).

## Discussion

This study is the first to present associations between IgG *N*-glycosylation and prevalent as well as incident vascular complications of type 2 diabetes, in three large cohort studies. In a meta-analysis, we found negative associations between galactosylation and sialylation and incident nephropathy during follow-up, also after adjustment for clinical risk factors. Furthermore, we found negative associations between galactosylation and sialylation and prevalent IHD. Galactosylation was also negatively associated with prevalent nephropathy, prevalent PAD and prevalent as well as incident retinopathy.

Type 2 diabetes and its complications result from complex interactions between environmental, metabolic and genetic factors and several of its risk factors are already known to influence IgG *N*-glycosylation patterns ^9^. IgG *N*-glycan patterns are partly heritable and have temporal stability ^20–22^, but can change in time by changes in cellular environment and disease, and have shown to influence IgG function ^3,23^. Associations between inflammation and all microvascular and macrovascular complications have been demonstrated. IgG glycans are therefore potential biomarkers for complications of type 2 diabetes.

### Microvascular complications

The pathogenesis of nephropathy is complex, multifactorial and yet to be fully elucidated. We now demonstrate that galactosylation and sialylation are negatively associated with both prevalent and incident diabetic nephropathy. It has been documented that hyperglycaemia in type 2 diabetes induces activation of inflammatory pathways, causes advanced glycation end-products, reactive oxygen species and activation of RAAS, eventually leading to tissue damage and vascular complications ^24–26^. Subsequent pro-inflammatory cascades lead to changes in kidney structure and function. The essential role of inflammation, and involvement of IgG was shown *in vivo* by Lopez-Parra et al. ^27^, demonstrating that Fcγ receptor blockade is renoprotective, reducing renal hypertrophy, inflammation, and fibrosis in diabetic mice. There is evidence to suggest that galactosylation and sialylation can alter the effector function of IgG. Studies have shown the relationship between low galactosylation in IgG *N*-glycans and increased disease severity in various auto-immune diseases, indicating the ability of galactose to decrease inflammation ^28^. Notably, nonclinical laboratory studies have found contradictory results with an increase of galactosylation induced inflammation through the alternative and lectin route of the complement system ^8^ or enhancement of the classical pathway of the complement cascade ^29^. These nonclinical laboratory findings are, however, not in line with findings in most clinical studies so far. Additionally, IgG is thought to acquire anti-inflammatory properties upon glycan sialylation as it potentially decreases binding affinities for Fcγ receptors and total IgG sialylation decreases during an active pro-inflammatory immune response ^30^. However, the exact biological mechanism remains incompletely understood to date. Thus, galactosylation and sialylation cannot be considered anti-inflammatory in all circumstances due to the complexity of biological pathways. The associations found in this study are in line with our previous work in the DiaGene Study where monogalactosylated structures and GP14 (FA2G2) were associated with slower kidney function decline ^31^. In a non-diabetic population, Barrios et al. ^7^ found that individuals with galactosylated (notably GP14) and sialylated IgG glycans had a lower risk of developing chronic kidney disease. Furthermore, another study showed that agalactosylated IgG is associated with a more rapid eGFR decline in type 1 diabetes ^32^.

Similarly in retinopathy, hyperglycaemia activates inflammatory processes leading to oxidative stress, vascular leakage and eventually ganglion cell loss in the retina ^33^. We found negative associations for galactosylation and bisection at baseline. Galactosylation was also negatively associated with retinopathy during follow-up in the basic model, which could indicate an indirect effect through clinical risk factors or merely lower power in the full model, as the HR was similar to the basic model. As discussed above, galactosylation is thought to have an anti-inflammatory effect on IgG. Conversely, *in vitro* studies have shown that bisecting GlcNAc increases FcγRIII affinity and results in a pro-inflammatory effect ^34,35^. The negative association with bisection therefore seems contradictory and cannot be fully explained. Recently, Wu et al. ^36^ found similar associations with diabetic retinopathy in a retrospective study. Whether this is an effect associated with an inflammatory response triggered in the presence of retinopathy is unknown. We did not see a similar association in the prospective analyses, making it difficult to determine whether the finding is cause or consequence, although our results suggest the latter.

### Macrovascular complications

For macrovascular complications, we found associations at baseline, but not at follow up nor in the more extensive models, which suggests they may be more reflective of the presence of the complication and its risk factors rather than causal involvement. IHD was negatively associated with sialylation. Moreover, both IHD and PAD were negatively associated with galactosylation. These findings again indicate a potential pro-inflammatory state of IgG associated with macrovascular disease. Atherosclerosis is the major cause of IHD and PAD and is a chronic inflammatory condition of the artery walls. However, we did not find any significant associations for IHD and PAD during follow-up and the baseline results did not remain significant after adjustment for clinical risk factors. The small number of events in both IHD and PAD groups might have led to insufficient statistical power. It is therefore possible we were unable to detect a true effect in this study. In addition, risk factors of macrovascular disease are known to influence IgG *N*-glycosylation patterns ^9,37,38^. Therefore, the associations found for IHD and PAD could (partly) be attributed to the influence of risk factors on IgG glycosylation. Recently, Menni et al. ^12^ described associations between IgG *N*-glycan patterns and 10-year atherosclerotic cardiovascular disease risk score in individuals without diabetes. Among the glycan traits associated with a low CVD risk score found by Menni et al. were GP14 and GP18, which contain galactose and sialic acid. Interestingly, these glycan peaks had a similar direction of effect as the corresponding glycan features in our prospective analyses, although these associations were not statistically significant.

We did not find any significant associations for IBD in meta-analysis. Recently, a cross-sectional study by Di Liu et al. ^39^ did find significant differences in IgG *N*-glycan patterns between ischemic stroke patients and healthy controls. However, in the present study, IBD includes hemorrhagic stroke, transient ischemic attack and ischemic stroke, and therefore entails a multitude of etiologies, which individually may or may not be linked to glycosylation. Additionally, in diabetes, different mechanisms contribute to atherogenesis compared to individuals without diabetes.

### Strengths and Limitations

A major strength of this study is the use of three large independent populations that represent both primary and secondary type 2 diabetes care, which allowed us to meta-analyse our findings and optimise statistical power. Furthermore, we had extensive phenotyping in all cohorts that allowed adjustment for known confounders of type 2 diabetes complications and known factors affecting IgG *N*-glycosylation. In addition, due to the prospective nature of our studies, we were able to indicate the temporal sequence between IgG *N*-glycan patterns and diabetic complications. For nephropathy and partly for retinopathy the findings suggest that IgG *N*-glycosylation patterns precede the onset of the complication. This is important in light of biomarker potential and gives us a glance at the potential role of IgG *N*-glycosylation in the pathophysiology of diabetes complications. Future experimental studies and Mendelian randomisation should be conducted to clarify this potential causation further. However, considering causality, it cannot be excluded that preclinical aspects of nephropathy were present at baseline that have driven the association rather than IgG *N*-glycosylation having a causal effect on future complication development.

In our study, IgG *N*-glycosylation was measured at baseline in all cohorts. It is known that glycosylation can change in case of acute inflammation ^40^, and that glycans remain fairly stable over time ^6^, nonetheless, multiple measurements during follow-up may have given a better insight into the impact of glycosylation on diabetic complications. Even though this is one of the largest studies to investigate the relationship between IgG *N*-glycans and vascular complications to date, macrovascular complications are still quite rare and thus underpowered, possibly explaining a lack of significant results in prospective analyses. Of note, 27 % of the participants of the three cohorts had more than one vascular complication at baseline. It is possible that the same IgG *N*-glycan patterns precede or succeed different vascular complications and may thereby confound each other’s association. This would however have led to an underestimation rather than an overestimation of the results. Due to limited statistical power, it was not possible to adjust for the presence of multiple complications.

## Conclusion

To conclude, in three large type 2 diabetes cohorts, we show that IgG *N*-glycosylation is significantly associated with incident nephropathy and retinopathy and with prevalent IHD and PAD. The IgG associated glycosylation features are in line with previous reports on IgG glycosylation in other health conditions where inflammation plays a key role. This fuels genetic, experimental and epidemiological studies to dive further into the underlying mechanisms and predictive potential of IgG *N*-glycosylation in the context of diabetes complications.

## Data Availability

All data produced in the present study are available upon reasonable request to the authors

## Abbreviations

DCS: Hoorn Diabetes Care Study;
GDM: GenoDiabMar;
IBD: Ischemic Brain Disease;
IHD: Ischemic Heart Disease;
Non*-*HDL-c: Cholesterol value minus HDL cholesterol;
PAD: Peripheral Artery Disease.

## Author contributions

E.M. developed automated method for glycomics study and performed glycomic analysis of the Hoorn DCS cohort, processed Hoorn DCS glycomics raw data, developed the R-scripts for data visualization, wrote and reviewed/edited the manuscript. R.H. completed the DiaGene database, performed statistical analyses, wrote and reviewed/edited the manuscript. R.C.S. performed statistical analyses and reviewed/edited the manuscript. A.S. contributed to the collection and design of the GenoDiabMar (GDM) study, reviewed/edited the manuscript. O.G. reviewed/edited the manuscript. J.W.J.B coordinates the DCS study and reviewed/edited the manuscript. M.H. performed glycomic analysis of the DiaGene cohort and reviewed/edited the manuscript. P.E. coordinates the DCS study and reviewed/edited the manuscript. J.P reviewed/edited the manuscript. E.S. contributed to the collection, design, and coordination of the DiaGene study, reviewed/edited the manuscript. G.L. contributed to the conception of research question, reviewed/edited the manuscript, and contributed to the discussion. V.D. supervised automated-method development for glycomics study in DCS cohort, developed R-script for MS data processing, interpreted results, reviewed/edited the manuscript, and contributed to the discussion. C.B. coordinated the GDM study, contributed to the conception of research question, reviewed/edited the manuscript, and contributed to the discussion. L.’t H. contributed to the coordination of the Hoorn DCS Study, performed statistical analyses, and reviewed/edited the manuscript. M.W. supervised glycomics study, contributed to discussion, and reviewed/edited the manuscript. M.v.H. contributed to the conception of research question, collection, and coordination of the DiaGene study, reviewed/edited the manuscript, and contributed to the discussion.

## Data availability

The datasets generated during and/or analyzed during the current study are available from the corresponding author upon reasonable request.

## Statements of assistance

We wish to thank Dr. Marco R. Bladergroen, Dr. Simone Nicolardi and Jan Nouta for their support with the mass spectrometry analyses. The authors thank the study participants and the research staff of the DiaGene Study Eindhoven, Hoorn DCS, and GDM Study Spain.

## Guarantor

M.v.H is the guarantor of this work and, as such, had full access to all the data in the study and takes responsibility for the integrity of the data and the accuracy of the data analysis.

## Funding

E.M. was supported by funding from the European Union’s Horizon 2020 research and innovation program under the Marie Sklodowska–Curie grant for the project GlySign (contract no. 722095). O.G. and G.L. were supported by the European Structural and Investment funding for the ‘Croatian National Centre of Research Excellence in Personalized Healthcare’ (contract #KK.01.1.1.01.0010), ‘Centre of Competences in Molecular Diagnostics’ (contract #KK.01.2.2.03.0006), and the European Regional Development Fund grant ‘CardioMetabolic’ agreement (#KK.01.2.1.02.0321). C.B. is funded by grants FIS-FEDER-ISCIII PI16/00620 (Ext 2021) and Strategic Plan for Research and Innovation in Health, CatSalut, PERIS STL008 (2019-2021) to develop clinical and epidemiological studies mainly focused in diabetes and its associations with new biomarkers. M.v.H. was supported by the ErasmusMC fellowship. The study funders were not involved in the design of the study; the collection, analysis, and interpretation of data; writing the report; and did not impose any restrictions regarding the publication of the report.

## Conflict of interest

M.W. is inventor on a patent application on sialic acid derivatization by ethyl esterification, and E.M. and M.H. are employed by Genos Ltd; V.D. is currently employed by Janssen Biologics BV. No other potential conflicts of interest relevant to this article were reported.

## Notes

### Competing Interest Statement

The authors have declared no competing interest.

### Author Declarations

The medical ethics committees of the Erasmus Medical Center, VU University Medical Center Amsterdam, and Institut Mar d Investigacions Mediques have approved DiaGene (DiaGene MEC 2004-230), Hoorn DCS (Hoorn DCS MEC 2007-57), and GenoDiabMar (MEC 2014/4323) studies, respectively. All individuals gave their written informed consent

