## supplementary figures and methods for "IgG *N-*glycans are associated with prevalent and incident complications of type 2 diabetes"

Short running title:

*IgG glycans and type 2 diabetes complications*

Elham Memarian [a,b *], Ralph Heijmans [c *], Roderick C. Slieker [d,e], Adriana Sierra [f], Olga Gornik [g], Joline WJ Beulens [e,h], Maja Hanic [b], Petra Elders [i], Julio Pascual [f], Eric Sijbrands [c], Gordan Lauc [b,g], Viktoria Dotz [a **,^], Clara Barrios [f ** @], Leen M 't Hart [d,e,j **], Manfred Wuhrer [a #], Mandy van Hoek [c # @]

[a] Center for Proteomics and Metabolomics, Leiden University Medical Center, the Netherlands

[b] Genos Glycoscience Research Laboratory, Zagreb, Croatia

[c] Department of Internal Medicine, Erasmus MC - University Medical Center, Rotterdam, the Netherlands

[d] Department of Cell and Chemical Biology, Leiden University Medical Center, the Netherlands

[e] Department of Epidemiology and Data Science, Amsterdam University Medical Center, location VUMC, Amsterdam Public Health institute, Amsterdam, The Netherlands

[f] Department of Nephrology, Hospital del Mar, Institut Mar d´Investigacions Mediques, Barcelona, Spain

[g] Faculty of Pharmacy and Biochemistry, University of Zagreb, Zagreb, Croatia

[h] Julius Center for Health Sciences and Primary Care, University Medical Center Utrecht, Utrecht, the Netherlands

[i] Department of General Practice, Amsterdam Public Health Institute, Amsterdam UMC, location VUmc, Amsterdam, the Netherlands

[j] Department of Biomedical Data Sciences, Section Molecular Epidemiology, Leiden University Medical Center, the Netherlands

[*] Shared first

[**] Shared penultimate

[#] Shared last

[@] Shared corresponding

[^] Present address: BioTherapeutics Analytical Development, Janssen Biologics BV, Einsteinweg 101, 2333 CB Leiden, The Netherlands

**Corresponding authors:**

Mandy van Hoek (ORCIDID: 0000-0002-2957-5436/ Department of Internal Medicine, Erasmus MC – University Medical Center Rotterdam, Doctor Molewaterplein 40, 3015 GD, Rotterdam, The Netherlands/ Tel: 0031 633343603/)

Clara Barrios, (Department of Nephrology, Hospital del Mar, Institut Mar d'Investigacions Mediques, Passeig Marítim de la Barceloneta 25, 08003, Barcelona, Spain /Tel: 0034 932 48 31 62/)

### Supplementary Figures

**Supplementary Figure 1:**

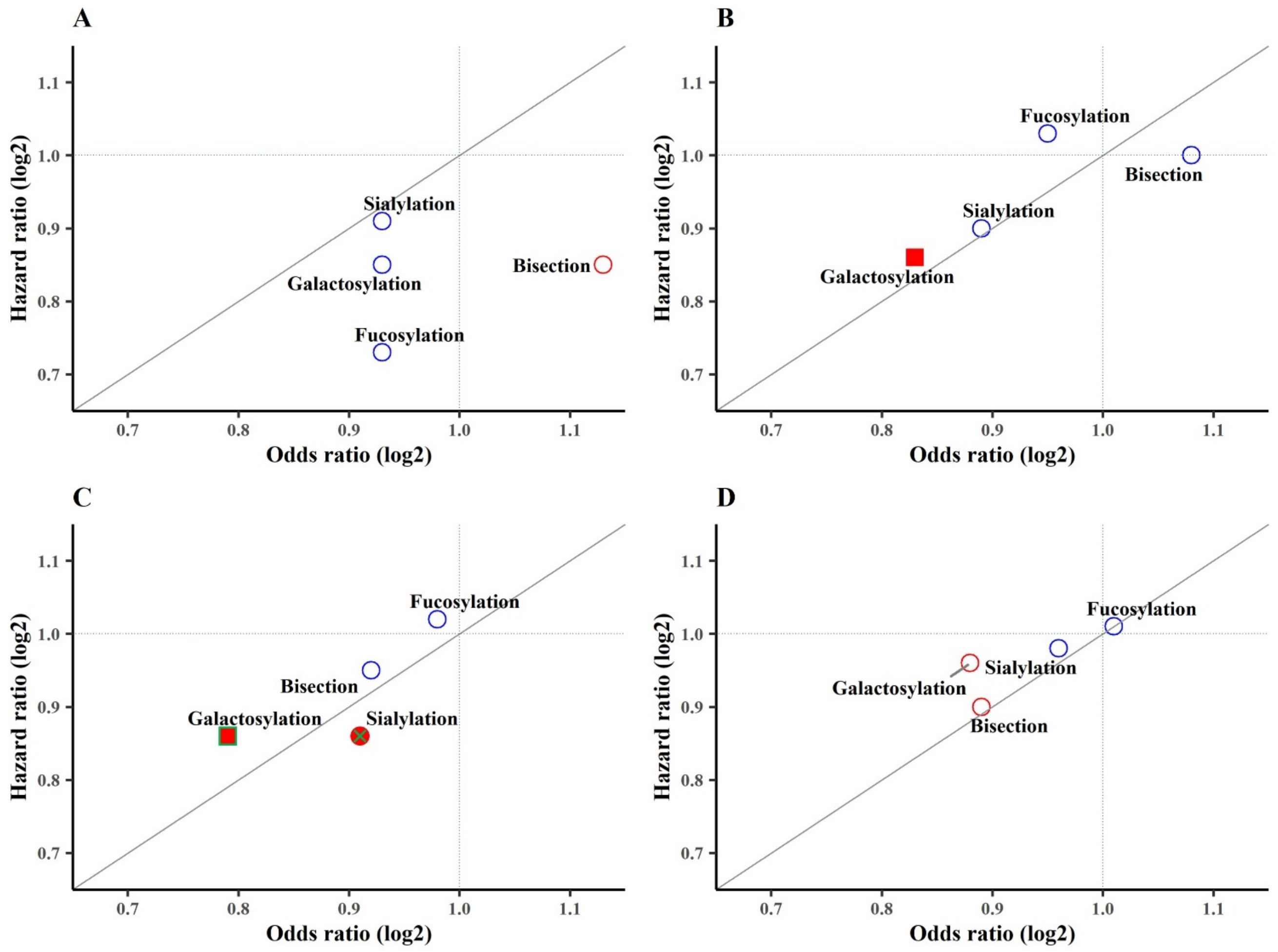


*N*-glycan derived traits odds ratios (OR) and hazard ratios (HR) for meta-analyzed data from DiaGene, Hoorn DCS, and GenoDiabMar studies in the full model (adjusted for age, sex, the interaction thereof, BMI, HDL, non-HDL, duration of diabetes, eGFR and HbA1c).

(a) Ischemic Heart Disease, (b) Peripheral Artery Disease, (c) Nephropathy, (d) Retinopathy. Red-filled green square: Significant in prevalent and incident complications after FDR correction. Red unfilled square with green cross: Significant in prevalent complications before FDR correction and in incident complications after FDR correction. Red-filled square: Significant in prevalent complications after FDR correction. Green-filled circle: Significant in incident complications before FDR correction. Red-filled circle with green cross: Significant in incident complications after FDR correction. Red unfilled circle: Significant in prevalent complications before FDR correction. Blue unfilled circle: Non-significant.

**Supplementary Figure 2:**

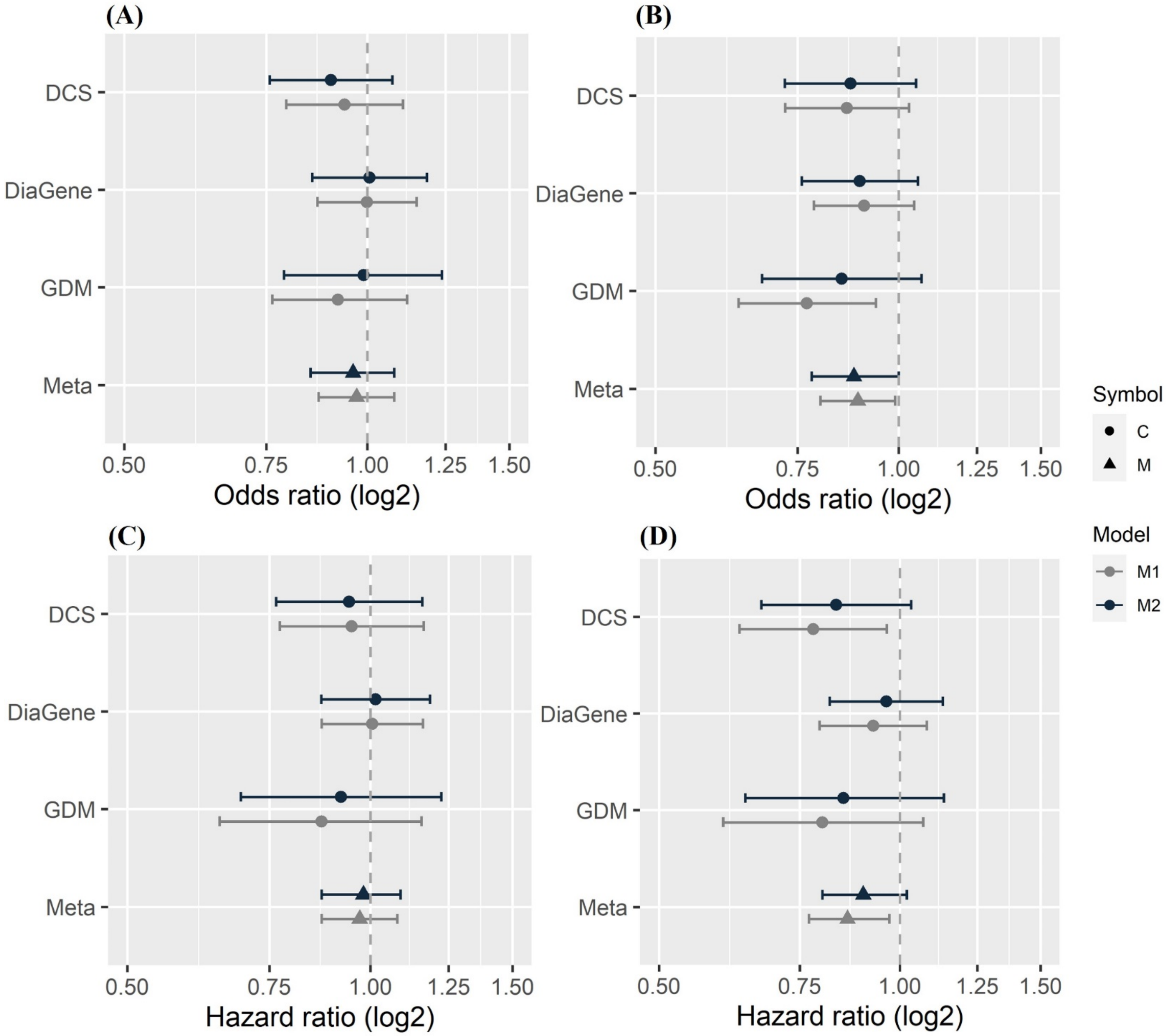


Forest plots of associations of *N*-glycan sialylation and galactosylation derived traits with retinopathy. Data from DiaGene, Hoorn DCS, and GenoDiabMar (GDM) studies and meta-analyzed data are from the basic and full model analysis.

(a) IgG Sialylation in Retinopathy (prevalent), (b) IgG Galactosylation in Retinopathy (prevalent), (c) IgG Sialylation in Retinopathy (incident), (d) IgG Galactosylation in Retinopathy (incident).

**Supplementary Figure 3:**

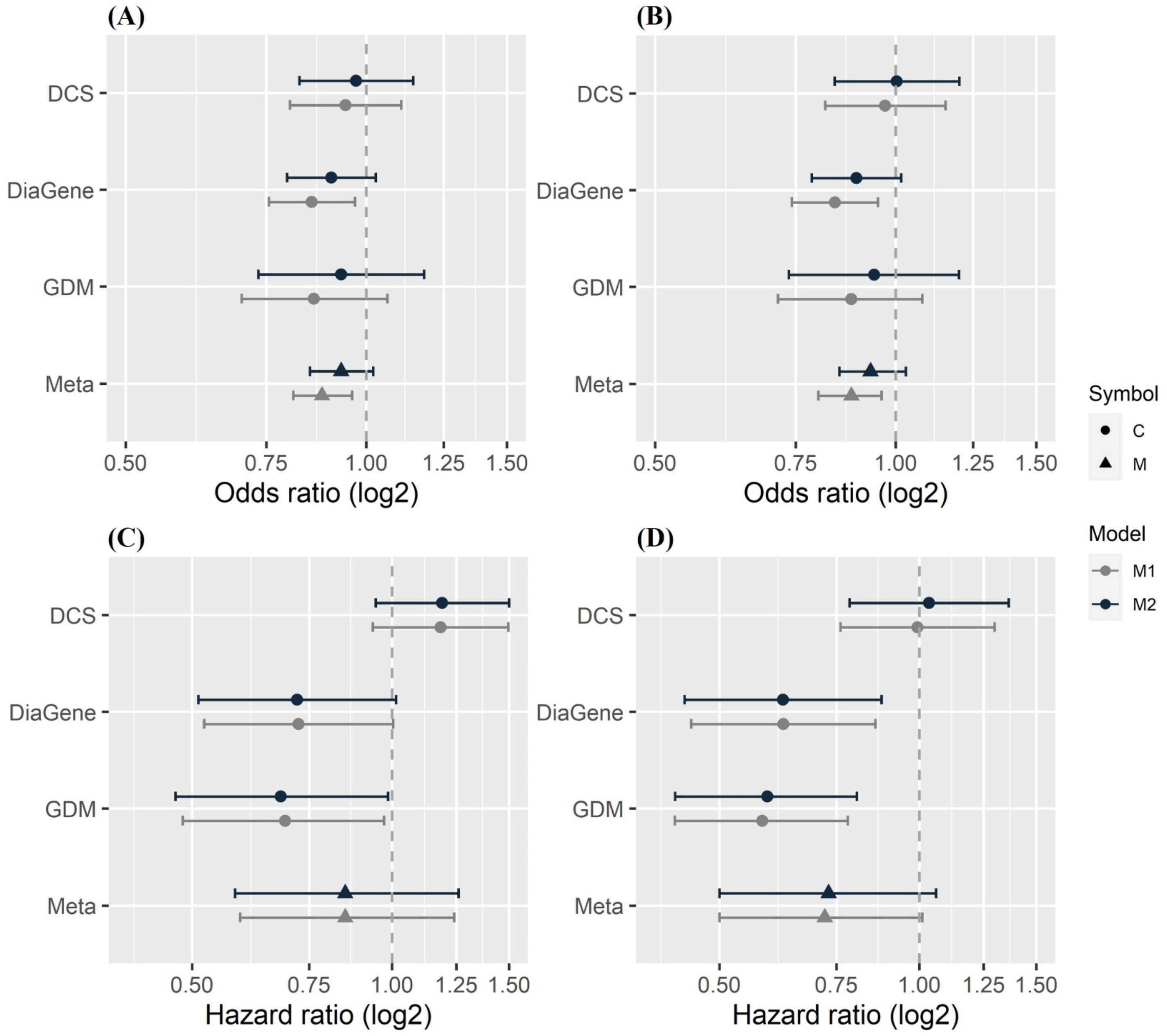


Forest plots of associations of *N*-glycan sialylation and galactosylation derived traits with ischemic heart disease (IHD). Data from DiaGene, Hoorn DCS, and GenoDiabMar (GDM) studies and meta-analyzed data are from the basic and full model analysis.

(a) IgG Sialylation in IHD (prevalent), (b) IgG Galactosylation in IHD (prevalent), (c) IgG Sialylation in IHD (incident), (d) IgG Galactosylation in IHD (incident).

**Supplementary Figure 4:**

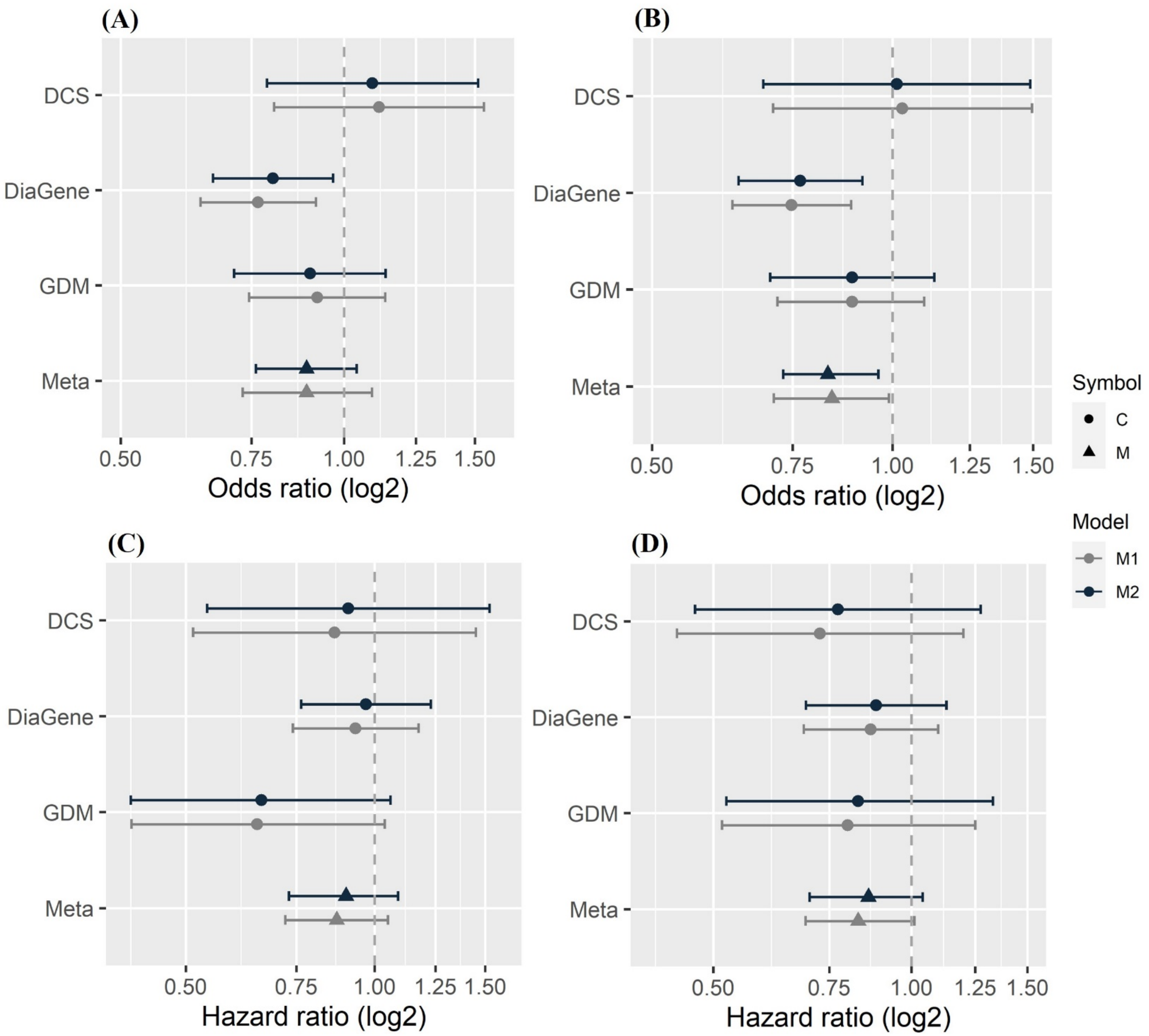


Forest plots of associations of *N*-glycan sialylation and galactosylation derived traits with peripheral artery disease (PAD). Data from DiaGene, Hoorn DCS, and GenoDiabMar (GDM) studies and meta-analyzed data are from the basic and full model analysis.

(a) IgG Sialylation in PAD (prevalent), (b) IgG Galactosylation in PAD (prevalent), (c) IgG Sialylation in PAD (incident), (d) IgG Galactosylation in PAD (incident).

#### Abbreviations

EDC: 1-ethyl-3-(3-dimethylaminopropyl) carbodiimide; HOBt: 1-hydroxybenzotriazole; MALDI-FTICR-MS: matrix-assisted laser desorption; ionization Fourier transform ion cyclotron resonance mass spectrometry; PNGase F: peptide-N-glycosidase F; SDS: sodium dodecyl sulfate; RMP: revolutions per minute; TEA: triethylamine; ACN: acetonitrile.

#### Materials and reagents

Analytical grade ethanol, sodium dodecyl sulfate (SDS), and trifluoroacetic acid (TFA) was supplied by Merck (Darmstadt, Germany). Sodium chloride (NaCl), 85% phosphoric acid (H3PO4), 50% sodium hydroxide (NaOH), nonidet P-40 substitute (NP-40), 1-hydroxybenzotriazole 97% (HOBt), and super-DHB (9:1 mixture of 2,5-dihydroxybenzoic acid and 2-hydroxy-5- methoxybenzoic acid, sDHB) were purchased from SigmaAldrich (Steinheim, Germany). 1-Ethyl-3-(3-(dimethylamino)-propyl) carbodiimide (EDC) hydrochloride was acquired from Fluorochem (Hadfield, UK), recombinant peptide-Nglycosidase F (PNGase F) was procured from Roche Diagnostics (Mannheim, Germany), and HPLC-grade acetonitrile (ACN) from Biosolve (Valkenswaard, The Netherlands). Milli-Q water (MQ) was obtained from an ELGA system (city, country) which was maintained at ≥18 MΩ. Disodium hydrogen phosphate dihydrate (Na2HPO4 × 2 H2O), potassium dihydrogen phosphate (KH2PO4), potassium chloride (KCl), and formic acid (FA), Protein G Sepharose® 4 Fast Flow (ProtG beads) and Orochem filterplates were purchased from SigmaAldrich (Steinheim, Germany).

**IgG capturing**

Human plasma samples (2 μL) were pre-diluted in 100 μL of 1×PBS. Using repetition pipette, 50 μL of ProtG beads slurry (beads + 1×PBS) was transferred per each well. After washing the beads three times with 200 μL 1×PBS on a vacuum manifold, 100 μL of the plasma-PBS mixture was added to each well followed by a 1 h incubation on a shaker (±1000 rpm). Subsequently, beads were washed three times with 200 μL 1×PBS and three times with 200 μL MQ on a vacuum manifold. After last round of washing, a plastic lid was placed on the filter plate. The filter plate was taken from the vacuum manifold and gently tapped on a Kimwipes Lint-Free tissue to remove any residual liquid from the bottom of the filter. The filter plate was placed with a separator on a clean PCR plate (FrameStar 96, fully skirted,hot foiled (Lot No.: 180174/ITOTAT (960/C)). Then, 100 μl of 100 mM FA was added to each well followed by 5 min on a shaker (±1000 rpm). Subsequently, the plate was centrifuged for 2 min at 100 g to elute the samples. The eluted samples were dried by vacuum centrifugation for 2 h at 60°C. Samples were resuspended by adding 4 μL 1×PBS to each well followed by incubation on shaker (±1000 rpm) for 5 min. Subsequently, 8 μL of 2% SDS was added to each well and each plate was covered with a silicone-lined mat followed by 5 min incubation on a shaker (±1000 rpm) and 10 min incubation at 60°C. The plate was allowed to reach room temperature, and 8.4 μL release mixture was added to each sample (4 μL 4% NP- 40, 4 μL 100 mM phosphoric acid in 5xPBS and 0.4 μL PNGase F). The plate was covered with a silicone-lined mat and placed for 5 min on a shaker at maximum speed, followed by overnight incubation at 37°C.

**N-glycan preparation and mass spectrometric analysis**

After *N*-glycan release, samples were stored at -20°C until further processing on an automated liquid handling platform as described previously (1). For this, sample plates were thawed on a shaker (±500 rpm) for 1 h. Solutions for automated sample processing were prepared, including sDHB matrix (2.5 mg/mL in 99% ACN with 1 mM NaOH), ethyl esterification reagent (0.25 M EDC with 0.25 HOBt in ethanol), 85% ACN, 85% ACN with 1% TFA and poured into specific containers on the robot. Cotton HILIC tips, pipetting tips, and microtitration plates were placed on the robot (1,2). 10 µL of released glycan sample was added by the robot to 50 µL ethyl esterification reagent, and the plate was incubated for 1 h at 37°C. Then, 50 µL of acetonitrile was added and after 10 min the solid phase extraction was started. For this, cotton HILIC tips were prewetted (3 x 40 µL of MQ water), and were conditioned (3 x 40 µL 85% ACN). Subsequently, samples were loaded (20 x up and down, 40 µL per time) followed by 2 washing steps (3 x 40 µL of 85% ACN containing 1% TFA) and washing of tips (3 x with 40 µL of 85% ACN), elution was done in MQ water (5 x pipetting up and down using 20 uL MQ water). Finally, 10 µL of purified samples were premixed with 5 µL of sDHB matrix and 6 µL of this mixture was spotted on a MALDI target plate (800/384 MTP AnchorChip, Bruker Daltonics). The spots were allowed to air-dry, followed by MALDI-FTICR-MS measurement as described previously (2). MS data were processed and subjected to quality control as previously described (3).
